# Agreement between DSM-IV and DSM-5 measures of substance use disorders in a sample of adult substance users

**DOI:** 10.1101/2020.10.26.20219840

**Authors:** Ofir Livne, Malka Stohl, Dvora Shmulewitz, Zachary Mannes, Deborah Hasin

**Author notes:** **Corresponding Author** Deborah S. Hasin, Ph.D., Columbia University/New York State, Psychiatric Institute 1051 Riverside Drive, Box, 123, New York, NY 10032.

## Abstract

**Aim:** In DSM-5, definitions of substance use disorders (SUD) were changed considerably from DSM-IV, yet little is known about how well DSM-IV and DSM-5 SUD diagnoses agree with each other within a series of individuals with substance use problems.

**Methods:** Prevalences and chance-corrected agreement of DSM-5 SUD and DSM-IV substance dependence were evaluated in 588 adult substance users, interviewed by clinician interviewers using the semi-structured Psychiatric Research Interview for Substance and Mental Disorders (PRISM-5). Alcohol, tobacco, cannabis, cocaine, heroin, opioid, sedative, and stimulant use disorders were examined. Cohen’s kappa was used to assess agreement between DSM-5 and DSM-IV SUD (including abuse or dependence), DSM-5 SUD and DSM-IV dependence, and DSM-5 moderate-to-severe SUD and DSM-IV dependence.

**Results:** Agreement between DSM-5 and DSM-IV SUD was excellent for alcohol, cocaine, heroin, opioids, sedatives, and stimulants (*κ*=0.84; 0.91; 0.99; 0.96; 0.92; 0.97; respectively) and substantial for alcohol and tobacco *(κ*=0.75; 0.80, respectively). Agreement between DSM-5 SUD and DSM-IV substance dependence was excellent for cocaine, heroin, opioids, sedatives, and stimulants (*κ*=0.89; 0.97; 0.90; 0.88; 0.94, respectively) and substantial for alcohol, tobacco, and cannabis *(κ*=0.75; 0.69; 0.63, respectively). Agreement between moderate and severe DSM-5 SUD and DSM-IV dependence was excellent across all substances.

**Conclusion:** Findings suggest that while care should always be used in interpreting the results of studies using different methods, studies relying on DSM-IV or DSM-5 SUD diagnostic criteria can be considered to offer similar information and thus can be compared when accumulating a body of evidence on a particular issue regarding substance use.

## INTRODUCTION

Substance use and substance use disorders (SUD) are a leading cause of morbidity and mortality [1]. Adults diagnosed with SUD are at increased risk of impaired functioning, psychiatric comorbidity, and poverty [2-4]. Considering the burden of disease associated with SUD in the US and globally, substance use remains a serious public health concern [5]. Reliable and valid measures of SUD are an essential component of research to successfully identify etiologic factors and effective treatments for SUD and associated negative outcomes.

In 1980, the third edition of the Diagnostic and Statistical Manual of Mental Disorders (DSM-III) was published. This was the first U.S. nomenclature to base diagnoses of substance and mental disorders on specific diagnostic criteria. DSM-III divided SUD into two disorders, abuse and dependence. Since then, successive versions of DSM have become the standard classification reference for clinical, research, policy, and reimbursement in the US and in many other countries [6-8]. This includes DSM-III-R (revised) in 1987 [9], which reorganized the SUD criteria into a concept of substance dependence [10] developed through empirical clinical research and recommended by the World Health Organization [11], and a residual category for abuse. DSM-IV[12], the next iteration, was published in 1994, and largely maintained the DSM-III-R SUD categories of dependence and abuse. Many studies supported the reliability and validity of the dependence diagnosis, but raised questions about the reliability and validity of the abuse category [13] and problems due to “diagnostic orphans”, i.e., those with two dependence criteria but who failed to receive a DSM-IV substance use disorder diagnosis because they had no abuse criteria and did not meet the dependence threshold of 3 or more criteria [14, 15].

After publication of DSM-IV, scientific knowledge about substance use disorders continued to grow, including substantial evidence indicating psychometric problems created by the hierarchical distinction between the DSM-III-R and DSM-IV abuse and dependence distinction, and other studies showing a unidimensional structure of the abuse and dependence criteria [16]. This led to recommendations to replace the abuse and dependence diagnoses with one combined SUD diagnosis [13, 17-19]. Consequently, major changes were made to the DSM-5 criteria for SUD [16], the most recent version of DSM, published in 2013. The most significant changes included: 1) merging the criteria of substance abuse and substance dependence into one all-encompassing disorder; 2) requiring two criteria to be met for the diagnosis of SUD, as opposed to the DSM-IV requirements that three criteria be met for the diagnosis of dependence and one criterion be met for the diagnosis of abuse; 3) adding craving – a strong desire or urge to use the substance – as a diagnostic criterion; 4) adding cannabis withdrawal as a new disorder to the list of other withdrawal syndromes already found in DSM-III-R and DSM-IV, and as a criterion for DSM-5 cannabis use disorder. These changes mainly aimed to overcome DSM-IV limitations, specifically, the lack of consistency between abuse and dependence in terms of number of criteria for diagnosis, and the low reliability and validity of substance abuse, compared to substance dependence, which was consistently shown to be a highly reliable [20] and valid syndrome [21].

Since its publication, the DSM-5 SUD diagnostic criteria have been widely adopted by clinicians and researchers to assess risk factors and consequences of substance use, and to identify participants for inclusion in clinical trials of treatment effectiveness. Nevertheless, a wide variety of researchers continue to rely on DSM-IV diagnoses because data from many studies, including large clinical trials [18, 22-26] and U.S yearly national general population surveys are based on DSM-IV diagnoses of SUD [27].

Recently, test-retest reliability of DSM-5 SUD diagnosis and severity levels (mild, moderate and severe disorder; 2–3, 4–5, and ≥6 criteria, respectively) were shown to have substantial to excellent reliability for most substances [28]. Nevertheless, few studies have empirically assessed the agreement of DSM-5 SUD and its severity levels with DSM-IV abuse and dependence diagnoses. The DSM-5 work group concluded that using a threshold of two or more criteria to diagnose SUD would maximize agreement on the prevalence of DSM-IV substance abuse and dependence disorders combined [8]. However, since then, only two studies have examined agreement between DSM-5 and DSM-IV diagnoses in adult clinical samples [29, 30]. These studies were limited in that they assessed only lifetime and not current SUD diagnoses. Further, one of these studies focused only on alcohol use disorder (AUD) and used a subpopulation of prison inmates [30].

Given the widespread use of DSM-5 SUD diagnoses in many studies, albeit with continuing use of DSM-IV in other large-scale studies, more information is needed on the extent to which diagnoses based on the DSM-5 diagnostic criteria are concordant with diagnoses based on DSM-IV dependence. In particular, since the evidence for the reliability and validity of substance dependence was much stronger than the evidence for substance abuse, information is needed on whether the low threshold for a DSM-5 SUD diagnosis (2 criteria) leads to inclusion of cases who would formerly have received an abuse diagnosis, forming an overly heterogenous group of individuals with less validity than a group formed by using a higher threshold more analogous to the DSM-IV dependence diagnosis. Therefore, using the Psychiatric Research Interview for Substance and Mental Disorders (PRISM-5), which has been shown to be a reliable instrument to measure DSM-5 SUD [28], the current study aimed to determine the agreement between: 1) DSM-5 and DSM-IV past-year substance use disorder diagnoses; 2) DSM-5 past-year substance use disorder and DSM-IV past-year dependence diagnoses; and 3) DSM-5 past-year moderate or severe substance use disorder and DSM-IV past-year dependence diagnoses.

## METHODS

### Study Population and procedures

Details of the study and sample are provided elsewhere [28] and summarized here in brief. Study participants were adults aged 18 years or older recruited from a suburban inpatient addiction program and also, via newspaper and social media advertising, from an urban medical center. To be eligible, participants needed to report substance use in the prior 30 days or the 30 days prior to inpatient admission and endorse at least one DSM-5 substance use disorder criteria in pre-study screening. Of those who met pre-screening eligibility, on-site research coordinators described the study, screened further for eligibility and obtained informed consent from eligible participants. Baseline interviews were conducted with 588 participants (150 inpatients; 438 community participants) that constituted the analytic sample. Participants received $50 for the interview. All interviews were conducted between 05/11/2016 and 06/17/2019. Procedures were approved by Institutional Review Boards of New York State Psychiatric Institute and South Oaks Hospital.

### Diagnostic interview

The PRISM-5 interview is a semi-structured, computer-assisted interview designed for clinician interviewers, which covers the DSM-IV and DSM-5 symptoms and criteria of substance and psychiatric disorders. PRISM-5 differs from other diagnostic interviews by assessing substance disorders first, and by providing more detailed symptom data. Substances covered by the PRISM-5 include alcohol, cannabis, cocaine, hallucinogens, heroin, opioid painkillers, sedatives/tranquilizers, stimulants, and tobacco (cigarettes). In this study, we included all substances, except for hallucinogens, for which DSM-IV dependence and DSM-5 moderate/severe SUD showed low prevalence (≤2%).

### PRISM-5 substance screening

The substance disorder module begins with screening questions regarding use of each substance 6 or more times within any 12-month period. For substances available for medical use, screening asked about non-medical use (without a prescription or other than prescribed, for example, to get high). Among those screened positive, the SUD criteria are assessed.

### PRISM-5 substance use disorder measures

For each substance, the PRISM-5 asked participants about using the substance at least 6 times within any 12-month period. For substances available for medical use, the screening questions ask about non-medical use (without a prescription or other than prescribed). Among those answering positively to this screening question for each substance, the substance disorder diagnostic criteria were assessed. For each substance, current DSM-5 SUD was considered positive if participants endorsed ≥2 criteria of the 11 criteria in the past 12 months. A second variable indicated the DSM-5 SUD severity measure: no disorder (0-1 criteria), mild (2–3 criteria), moderate (4–5 criteria), or severe (≥6 criteria). A third DSM-5 variable was constructed, which was positive for those with moderate or severe DSM-5 SUD and negative for those with no or mild disorder. DSM-IV current substance dependence was diagnosed when ≥3 of the DSM-IV criteria were endorsed within the past 12 months. A second binary variable was constructed to indicate DSM-IV substance use disorder, coded positive if participants endorsed dependence or abuse (based on endorsing ≥1 abuse criteria in the past 12 months).

### Sociodemographic variables

These were collected in an introductory PRISM-5 module. They included: sex (male; female), race/ethnicity (White; Black; Hispanic; other), age (18-29; 30-39; 40-49; 50+), marital status (not married; living together/married), education (high-school diploma or less; some college or more), employment (no job; any job).

### Interviewers, training and supervision

All interviewers had at least a master’s degree in psychology or social work and an average of 4.5 years of clinical experience (range, 1-10 years). PRISM-5 training included a manual, 2-day workshop, practice interviewing, role-playing, and certification. To become certified, trainees recorded mock interviews that underwent structured review by PRISM trainers. Trainees became interviewers after 2 recordings were rated satisfactory or better. Supervision of the study interviewers was conducted by trained, highly experienced trainer/supervisors with clinical masters degrees (psychology or social work) whose mean years of clinical experience utilizing the PRISM interview in research settings was 7.6 years (range: 3-10), and whose mean years of supervisory experience with the PRISM-5 was 6 years (range: 2-8). After certification, PRISM-5 supervisors met weekly with interviewers. They also reviewed recordings of 10% of the interviews, scoring interviewer performance, which occasionally indicated typical issues in such interviewing that required supervision (e.g., reading probes as written) but generally indicated that PRISM-5 interviews were largely conducted in a standardized way according to the PRISM-5 training procedures. Further information about the PRISM-5 and procedures used in this study can be found elsewhere [28].

### Analysis

Three analyses were conducted to determine the degree of agreement between substance-specific past-year DSM-IV and DSM-5 SUD measures. The first analysis compared DSM-5 with DSM-IV substance use disorder diagnoses. The second compared DSM-5 substance use disorder with DSM-IV past-year substance dependence diagnoses. The third compared DSM-5 moderate or severe substance use disorder with DSM-IV past-year substance dependence diagnoses. McNemar’s test evaluated whether prevalences differed between the DSM-IV and DSM-5 measure. Cohen’s kappa, a statistical measurement of chance-corrected agreement [31], was calculated for each analysis. The following standard interpretations were used for the degree of agreement indicated by kappa values: 0–0.20, poor; 0.21–0.40, fair; 0.41–0.60, moderate; 0.61–0.80, substantial; and 0.81–1.00, excellent [32]. All analyses were conducted using SAS 9.4 [33].

## RESULTS

### Sociodemographic characteristics of the sample

Respondents were primarily male (70%), black (42%), ages 50 years or above (50%), high school diploma or less (55%), not married (80%), and without a job (74%) (Table 1).

**Table 1:**
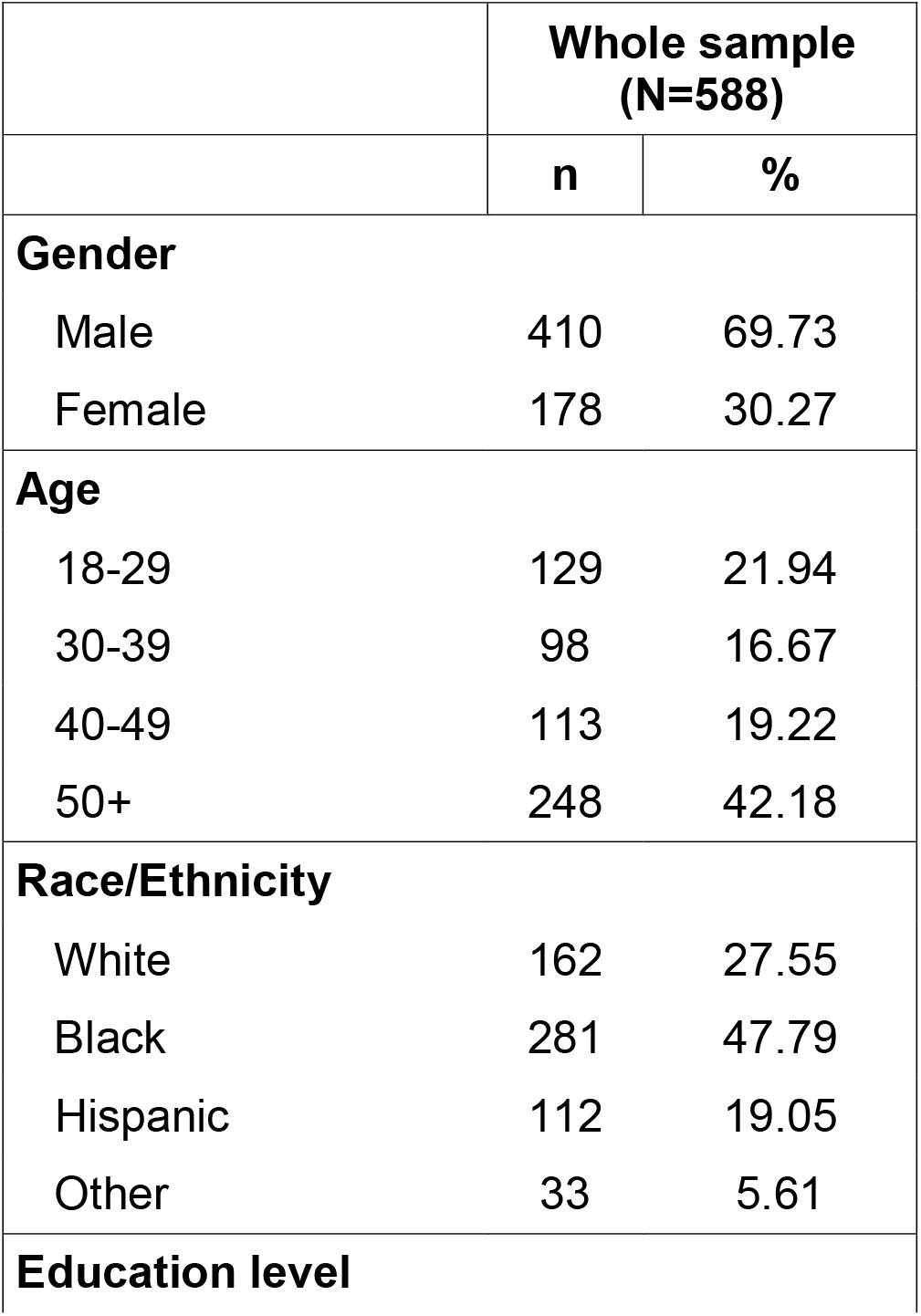
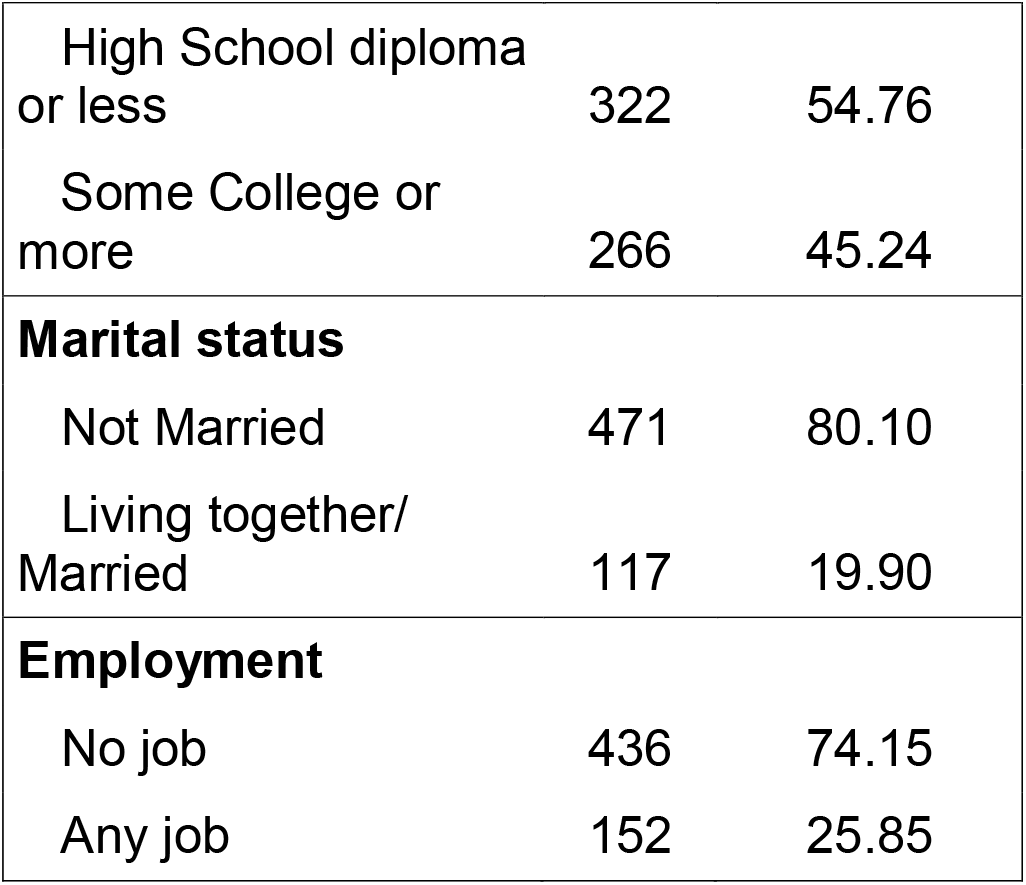
Sample descriptives, overall.

|  | Whole sample<br>(N=588) |  |
| --- | --- | --- |
|  | n | % |
| **Gender** |  |  |
| Male | 410 | 69.73 |
| Female | 178 | 30.27 |
| **Age** |  |  |
| 18-29 | 129 | 21.94 |
| 30-39 | 98 | 16.67 |
| 40-49 | 113 | 19.22 |
| 50+ | 248 | 42.18 |
| **Race/Ethnicity** |  |  |
| White | 162 | 27.55 |
| Black | 281 | 47.79 |
| Hispanic | 112 | 19.05 |
| Other | 33 | 5.61 |
| **Education level** |  |  |

|  | **Whole sample**<br>**(N=588)** |  |
| --- | --- | --- |
|  | **n** | **%** |
| High School diploma or less | 322 | 54.76 |
| Some College or more | 266 | 45.24 |
| **Marital status** |  |  |
| Not Married | 471 | 80.10 |
| Living together/<br>Married | 117 | 19.90 |
| **Employment** |  |  |
| No job | 436 | 74.15 |
| Any job | 152 | 25.85 |

### DSM-5 and DSM-IV substance use disorder

Table 2 shows prevalences of DSM-5 SUD and DSM-IV SUD (indicating a diagnosis of dependence or abuse). In DSM-5, prevalences of AUD, TUD and CUD were 66.0%, 62.1%, 44.6%, while corresponding prevalences in DSM-IV for these disorders were 59.4%, 52.2% and 33.2%. DSM-5 prevalences of cocaine, heroin and prescription opioid use disorders were 44.6%, 24.1% and 15.6%, while DSM-IV prevalences were 40.5%, 23.6% and 14.6%. Prevalences between DSM-5 SUD and DSM-IV SUD were significantly different for all substances, except for heroin and stimulants. Agreement between DSM-5 and DSM-IV SUD was excellent for alcohol, cocaine, heroin, opioids, sedatives, and stimulants (*κ*=0.84; 0.91; 0.99; 0.96; 0.92; 0.97, respectively) and substantial for alcohol and tobacco *(κ*=0.75; 0.80, respectively; Table 2).

**Table 2.**
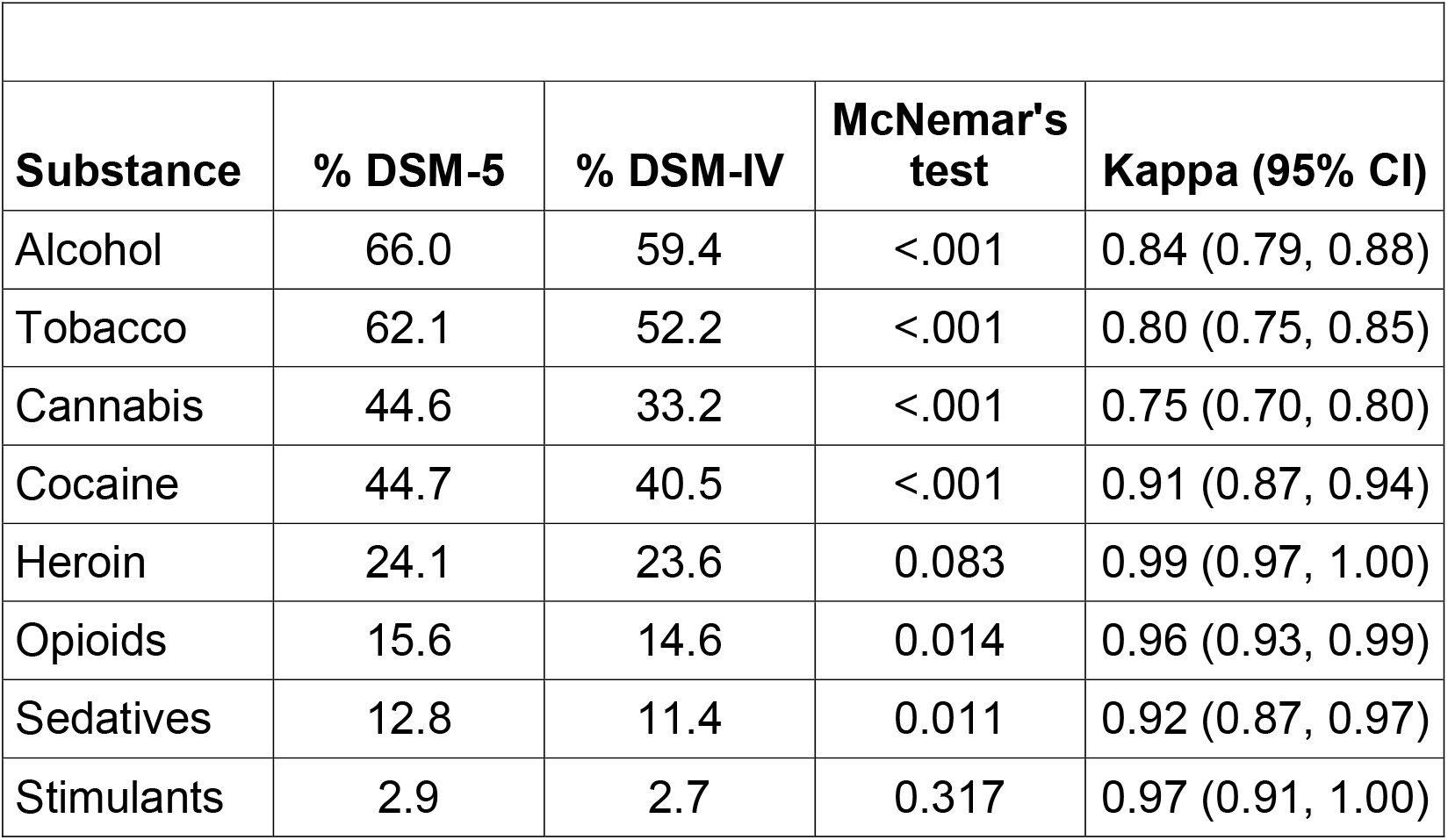
Agreement between PRISM-5 DSM-5 and DSM-IV past-year substance use disorder diagnoses (N=588)

| **Substance** | **% DSM-5** | **% DSM-IV** | **McNemar's test** | **Kappa (95% CI)** |
| --- | --- | --- | --- | --- |
| Alcohol | 66.0 | 59.4 | <.001 | 0.84 (0.79, 0.88) |
| Tobacco | 62.1 | 52.2 | <.001 | 0.80 (0.75, 0.85) |
| Cannabis | 44.6 | 33.2 | <.001 | 0.75 (0.70, 0.80) |
| Cocaine | 44.7 | 40.5 | <.001 | 0.91 (0.87, 0.94) |
| Heroin | 24.1 | 23.6 | 0.083 | 0.99 (0.97, 1.00) |
| Opioids | 15.6 | 14.6 | 0.014 | 0.96 (0.93, 0.99) |
| Sedatives | 12.8 | 11.4 | 0.011 | 0.92 (0.87, 0.97) |
| Stimulants | 2.9 | 2.7 | 0.317 | 0.97 (0.91, 1.00) |

### DSM-5 substance use disorder and DSM-IV dependence

Table 3 shows prevalences of DSM-5 SUD and DSM-IV substance dependence. The prevalences of DSM-IV dependence diagnoses were 59.4%, 52.2%, 33.2%, 40.5%, 23.6% and 14.6% for alcohol, tobacco, cannabis, cocaine, heroin, and prescription opioid dependence, respectively (Table 3). Agreement between DSM-5 SUD and DSM-IV substance dependence was excellent for cocaine, heroin, opioids, sedatives, and stimulants (*κ*=0.89; 0.97; 0.90; 0.88;0.94, respectively) and substantial for alcohol, tobacco, and cannabis *(κ*=0.75; 0.69; 0.63, respectively; Table 3).

**Table 3.** Agreement between PRISM-5 DSM-5 past-year substance use disorder and DSM-IV past-year substance dependence diagnoses (N=588)

| **Substance** | **% DSM-5** | **% DSM-IV** | **McNemar's test** | **Kappa (95% CI)** |
| --- | --- | --- | --- | --- |
| Alcohol | 66.0 | 53.7 | <.001 | 0.75 (0.70, 0.80) |
| Tobacco | 62.1 | 46.4 | <.001 | 0.69 (0.64, 0.75) |
| Cannabis | 44.6 | 26.9 | <.001 | 0.63 (0.57, 0.69) |
| Cocaine | 44.7 | 39.1 | <.001 | 0.89 (0.85, 0.92) |
| Heroin | 24.1 | 23.0 | 0.008 | 0.97 (0.94, 0.99) |
| Opioids | 15.6 | 13.1 | <.001 | 0.90 (0.85, 0.95) |
| Sedatives | 12.8 | 10.4 | <.001 | 0.88 (0.82, 0.94) |
| Stimulants | 2.9 | 2.6 | 0.157 | 0.94 (0.85, 1.00) |

### DSM-5 moderate or severe substance use disorder and DSM-IV dependence

Table 4 shows prevalences of combined moderate and severe levels of DSM-5 SUD and DSM-IV substance dependence. Differences in prevalence of DSM-5 moderate-severe SUD and DSM-IV substance dependence were negligible for most substances, e.g., 47.4% vs. 46.4% for tobacco (Table 4). The only exception to this was cannabis use disorder, whose prevalence of moderate-severe DSM-5 disorders was 30.8% but 26.9% for DSM-IV dependence. Agreement between moderate and severe DSM-5 SUD and DSM-IV dependence diagnosis was excellent across all substances, and near-perfect for cocaine and heroin (*κ*=0.97; 0.98, respectively; Table 4).

**Table 4.** Agreement between PRISM-5 DSM-5 past-year moderate or severe substance use disorder and DSM-IV past-year substance dependence diagnoses (N=588)

| **Substance** | **% DSM-5** | **% DSM-IV** | **McNemar's test** | **Kappa (95% CI)** |
| --- | --- | --- | --- | --- |
| Alcohol | 53.7 | 53.7 | 1.000 | 0.92 (0.89, 0.96) |
| Tobacco | 47.4 | 46.4 | 0.180 | 0.93 (0.90, 0.96) |
| Cannabis | 30.8 | 26.9 | <.001 | 0.86 (0.81, 0.90) |
| Cocaine | 38.9 | 39.1 | 0.705 | 0.97 (0.96, 0.99) |

| Substance | % DSM-5 | % DSM-IV | McNemar's test | Kappa (95% CI) |
| --- | --- | --- | --- | --- |
| Heroin | 23.0 | 23.0 | 1.000 | 0.98 (0.96, 1.00) |
| Opioids | 12.6 | 13.1 | 0.317 | 0.93 (0.89, 0.98) |
| Sedatives | 10.4 | 10.4 | 1.000 | 0.96 (0.93, 1.00) |
| Stimulants | 2.4 | 2.6 | 0.317 | 0.96 (0.90, 1.00) |

## DISCUSSION

In recent years, the DSM-5 SUD diagnostic criteria have been extensively utilized by researchers and clinicians worldwide. However, the previous DSM-IV diagnostic system is still being used in numerous studies, for example, ongoing analyses of large existing genetic or clinical trial datasets, or in the yearly national surveys of the National Drug Use and Health Survey (NSDUH). Therefore, determining the level of agreement between DSM-5 and DSM-IV SUD diagnostic systems is important to inform investigators and others about whether results from studies using DSM-IV or DSM-5 SUD are reasonably comparable. To date, this information is largely missing. Using rigorous methods and drawing on a study with several hundred participants that utilized rigorous assessment procedures, we examined chance-corrected agreement between DSM-5 and DSM-IV SUD diagnoses for six commonly used substances. In all sets of comparisons and across all substances, substantial to excellent levels of agreement were found between DSM-5 and DSM-IV diagnoses.

In this study, the prevalence of DSM-5 SUD was slightly higher than the prevalence of DSM-IV SUD for all substances, with differences ranging from 4.2%-11.4%. These differences slightly vary from results from previous studies, which used a DSM-IV substance use disorder variable that was similar to the one used in this study (coded positive if participants endorsed dependence or abuse). These variations are likely related to the different samples used in those previous studies and this one; whereas the current study’s sample included substance users only, previous studies used samples that mixed substance users with non-substance users [29, 34-37]. For all substances, differences in prevalence between DSM-5 SUD and DSM-IV dependence were consistently greater than differences in prevalence between DSM-5 and DSM-IV SUD prevalence. This finding is not surprising; it stems from the lower two-symptom threshold of the DSM-5 SUD diagnosis, compared to the three-symptom threshold of DSM-IV dependence. Further, the higher prevalences of DSM-5 SUD is consistent with the possibility that the lower threshold of the DSM-5 diagnostic criteria is inclusive of individuals with two dependence symptoms but no abuse symptoms who lacked any SUD diagnosis in DSM-IV.

Regardless of diagnostic threshold or type of comparison, DSM-5 and DSM-IV SUD diagnoses had substantial to excellent degrees of agreement with each other for all substances, with kappas ranging from 0.63 to 0.99. To our knowledge, this study is the first to report agreement between DSM-5 and DSM-IV tobacco, prescription opioid, heroin, sedative, and stimulant use disorders (*κ*=0.80; 0.99; 0.96; 0.92; 0.97, respectively). These findings are important as they provide more evidence that the underlying construct of a substance use disorder has robust validity, as its degree of agreement with dependence remains strong even when SUD measures somewhat differ in their operationalization [38].

Differences in prevalences of DSM-5 moderate-severe SUD and DSM-IV dependence were negligible. Accordingly, agreement between DSM-IV and DSM-5 was stronger (near-perfect for many substances) for this set of comparison than for others. This finding suggests that these two diagnoses may indicate similar clinical conditions; investigators utilizing information from studies that employ one classification system or the other, can have confidence in the interchangeability of these studies’ results.

Questions could be raised about the research and clinical utility of the DSM-5 SUD mild severity level from the current study’s report of better agreement between DSM-5 moderate-severe SUD and dependence. Substance users that endorse criteria for mild SUD may be recognized less frequently by clinicians but may still benefit from interventions that are appropriate for their lower severity level, e.g., the SBIRT model (Screening, Brief Intervention, Referral to Treatment [39], which has been shown to be effective particularly for alcohol [40]. Longitudinal studies that examine whether brief preventive and therapeutic interventions in primary care settings mitigate the development of more severe SUD among individuals with mild DSM-5 SUD are warranted, as these may have important individual and public health consequences.

Across the comparisons performed in this study, agreement between DSM-IV and DSM-5 for CUD was slightly lower than agreement for other substances. This is most likely due to the fact that a cannabis withdrawal criterion was not included in DSM-IV, but was added to the diagnostic criteria of CUD in DSM-5.

Study limitations are noted. First, the sample included adults treated for substance use problems in inpatient settings and a community sample of adults recruited via newspaper and social media advertising. Studies that examine agreement for DSM-5 and DSM-IV SUD in adolescents, in general mental health and primary care settings, and in general population representative samples are warranted. Second, analyses did not include some substances, such as hallucinogens, as prevalences for these were low. Studies that report agreement between DSM-5 and DSM-IV SUD diagnoses for these substances are needed. Finally, the current study utilized self-reported measures of substance use, which could have led to reporting inaccuracies. Despite these limitations, this study had several important strengths. First, the sample used in this study was relatively large and diverse; it included substance users with a wide range of SUD severity levels and from various treatment settings. Second, it included an adequate representation of different substances with substantial burden of disease; Finally, it employed rigorous study methods and reliable measures.

## Conclusion

The current study assessed the agreement between DSM-5 and DSM-IV SUD, using several comparison models, in which measurements were operationalized differently. In all comparisons performed and across all substances, agreements between DSM-5 and DSM-IV diagnoses were substantial to excellent. Further, excellent agreement between moderate or severe levels of DSM-5 SUD and DSM-IV dependence, a highly reliable measure, indicate that these specific DSM-5 SUD severity levels had even better agreement with DSM-IV dependence measures than DSM-5 SUD measures that included mild cases. Substance use and SUD remain a major public health concern.Use of reliable and valid diagnostic measures of these disorders are crucial if studies are to generate informative results on etiology, prevention and treatment. This study suggests that while care should always be used in interpreting the results of studies using different methods, studies relying on DSM-IV or DSM-5 SUD diagnostic criteria can be considered to offer similar information and thus can be compared when accumulating a body of evidence on a particular issue regarding substance use disorders.

## Data Availability

This study was conducted as a part of a larger study investigating reliability and validity of substance use disorders. For information regarding the data, please contact

## REFERENCES

1. Schulte, M.T. and Y.I. Hser, Substance Use and Associated Health Conditions throughout the Lifespan. Public Health Rev, 2014. 35(2).

2. O’Brien, C.P., D.S. Charney, L. Lewis, J.W. Cornish, R.M. Post, G.E. Woody, J.K. Zubieta, J.C. Anthony, J.D. Blaine, C.L. Bowden, J.R. Calabrese, K. Carroll, T. Kosten, B. Rounsaville, A.R. Childress, D.W. Oslin, H.M. Pettinati, M.A. Davis, R. Demartino, R.E. Drake, M.F. Fleming, L. Fricks, A.H. Glassman, F.R. Levin, E.V. Nunes, R.L. Johnson, C. Jordan, R.C. Kessler, S.K. Laden, D.A. Regier, J.A. Renner, Jr., R.K. Ries, T. Sklar-Blake, and C. Weisner, Priority actions to improve the care of persons with co-occurring substance abuse and other mental disorders: a call to action. Biol Psychiatry, 2004. 56(10): p. 703–13.

3. Grant, B.F., R.B. Goldstein, T.D. Saha, S.P. Chou, J. Jung, H. Zhang, R.P. Pickering, W.J. Ruan, S.M. Smith, B. Huang, and D.S. Hasin, Epidemiology of DSM-5 Alcohol Use Disorder: Results From the National Epidemiologic Survey on Alcohol and Related Conditions III. JAMA Psychiatry, 2015. 72(8): p. 757–66.

4. Grant, B.F., T.D. Saha, W.J. Ruan, R.B. Goldstein, S.P. Chou, J. Jung, H. Zhang, S.M. Smith, R.P. Pickering, B. Huang, and D.S. Hasin, Epidemiology of DSM-5 Drug Use Disorder: Results From the National Epidemiologic Survey on Alcohol and Related Conditions-III. JAMA Psychiatry, 2016. 73(1): p. 39–47.

5. Alcohol, G.B.D. and C. Drug Use, The global burden of disease attributable to alcohol and drug use in 195 countries and territories, 1990-2016: a systematic analysis for the Global Burden of Disease Study 2016. Lancet Psychiatry, 2018. 5(12): p. 987–1012.

6. Kupfer, D.J., E.A. Kuhl, and D.A. Regier, DSM-5--the future arrived. JAMA, 2013. 309(16): p. 1691–2.

7. van Heugten-van der Kloet, D. and T. van Heugten, The classification of psychiatric disorders according to DSM-5 deserves an internationally standardized psychological test battery on symptom level. Front Psychol, 2015. 6: p. 1108.

8. American Psychiatric Publishing. Insurance Implications of DSM-5. 2013. https://bluecare.bcbst.com/forms/ProviderForms/Insurance_Implications_of_DSM-5.pdf.

9. American Psychiatric Association, Diagnostic and Statistical Manual of Mental Disorders (3rd ed., revised). 1987, Arlington, VA.

10. Edwards, G. and M.M. Gross, Alcohol dependence: provisional description of a clinical syndrome. Br Med J, 1976. 1(6017): p. 1058–61.

11. Edwards, G., A. Arif, and R. Hadgson, Nomenclature and classification of drug- and alcohol-related problems: a WHO Memorandum. Bull World Health Organ, 1981. 59(2): p. 225–42.

12. American Psychiatric Association, Diagnostic and Statistical Manual of Mental Disorders (4th ed.). 1994, Arlington, VA.

13. Shmulewitz, D., K. Keyes, C. Beseler, E. Aharonovich, C. Aivadyan, B. Spivak, and D. Hasin, The dimensionality of alcohol use disorders: results from Israel. Drug Alcohol Depend, 2010. 111(1-2): p. 146–54.

14. Pollock, N.K. and C.S. Martin, Diagnostic orphans: adolescents with alcohol symptom who do not qualify for DSM-IV abuse or dependence diagnoses. Am J Psychiatry, 1999. 156(6): p. 897–901.

15. Ray, L.A., R. Miranda, Jr., I. Chelminski, D. Young, and M. Zimmerman, Diagnostic orphans for alcohol use disorders in a treatment-seeking psychiatric sample. Drug Alcohol Depend, 2008. 96(1-2): p. 187–91.

16. Hasin, D.S., C.P. O’Brien, M. Auriacombe, G. Borges, K. Bucholz, A. Budney, W.M. Compton, T. Crowley, W. Ling, N.M. Petry, M. Schuckit, and B.F. Grant, DSM-5 criteria for substance use disorders: recommendations and rationale. Am J Psychiatry, 2013. 170(8): p. 834–51.

17. Hasin, D.S., M.C. Fenton, C. Beseler, J.Y. Park, and M.M. Wall, Analyses related to the development of DSM-5 criteria for substance use related disorders: 2. Proposed DSM-5 criteria for alcohol, cannabis, cocaine and heroin disorders in 663 substance abuse patients. Drug Alcohol Depend, 2012. 122(1-2): p. 28-37.

18. Hagman, B.T. and A.M. Cohn, Toward DSM-V: mapping the alcohol use disorder continuum in college students. Drug Alcohol Depend, 2011. 118(2-3): p. 202–8.

19. Saha, T.D., W.M. Compton, S.P. Chou, S. Smith, W.J. Ruan, B. Huang, R.P. Pickering, and B.F. Grant, Analyses related to the development of DSM-5 criteria for substance use related disorders: 1. Toward amphetamine, cocaine and prescription drug use disorder continua using Item Response Theory. Drug Alcohol Depend, 2012. 122(1-2): p. 38–46.

20. Hasin, D., M.L. Hatzenbuehler, K. Keyes, and E. Ogburn, Substance use disorders: Diagnostic and Statistical Manual of Mental Disorders, fourth edition (DSM-IV) and International Classification of Diseases, tenth edition (ICD-10). Addiction, 2006. 101 Suppl 1: p. 59–75.

21. Compton, W.M., Y.F. Thomas, F.S. Stinson, and B.F. Grant, Prevalence, correlates, disability, and comorbidity of DSM-IV drug abuse and dependence in the United States: results from the national epidemiologic survey on alcohol and related conditions. Arch Gen Psychiatry, 2007. 64(5): p. 566–76.

22. Anton, R.F., K.E. Voronin, S.W. Book, P.K. Latham, P.K. Randall, W.B. Glen, M. Hoffman, and J.P. Schacht, Opioid and Dopamine Genes Interact to Predict Naltrexone Response in a Randomized Alcohol Use Disorder Clinical Trial. Alcohol Clin Exp Res, 2020.

23. Williams, E.C., J.F. Bobb, A.K. Lee, E.J. Ludman, J.E. Richards, E.J. Hawkins, J.O. Merrill, A.J. Saxon, G.T. Lapham, T.E. Matson, L.J. Chavez, R. Caldeiro, D.M. Greenberg, D.R. Kivlahan, and K.A. Bradley, Effect of a Care Management Intervention on 12-Month Drinking Outcomes Among Patients With and Without DSM-IV Alcohol Dependence at Baseline. J Gen Intern Med, 2019.

24. Brick, L.A., L. Micalizzi, V.S. Knopik, and R.H.C. Palmer, Characterization of DSM-IV Opioid Dependence Among Individuals of European Ancestry. J Stud Alcohol Drugs, 2019. 80(3): p. 319–330.

25. Santaella-Tenorio, J., N.S. Levy, L.E. Segura, P.M. Mauro, and S.S. Martins, Cannabis use disorder among people using cannabis daily/almost daily in the United States, 2002-2016. Drug Alcohol Depend, 2019. 205: p. 107621.

26. Compton, W.M., B. Han, C.M. Jones, and C. Blanco, Cannabis use disorders among adults in the United States during a time of increasing use of cannabis. Drug Alcohol Depend, 2019. 204: p. 107468.

27. Center for Behavioral Health Statistics and Quality. Impact of the DSM-IV to DSM-5 Changes on the National Survey on Drug Use and Health. 2016. Rockville, MD. https://www.samhsa.gov/data/sites/default/files/NSDUH-DSM5ImpactAdultMI-2016.pdf

28. Hasin, D., D. Shmulewitz, M. Stohl, E. Greenstein, S. Roncone, E. Aharonovich, and M. Wall, Test-retest reliability of DSM-5 substance disorder measures as assessed with the PRISM-5, a clinician-administered diagnostic interview. Drug Alcohol Depend, 2020. 216: p. 108294.

29. Peer, K., L. Rennert, K.G. Lynch, L. Farrer, J. Gelernter, and H.R. Kranzler, Prevalence of DSM-IV and DSM-5 alcohol, cocaine, opioid, and cannabis use disorders in a largely substance dependent sample. Drug Alcohol Depend, 2013. 127(1-3): p. 215-9.

30. Kopak, A.M., A.V. Metze, and N.G. Hoffmann, Alcohol use disorder diagnoses in the criminal justice system: an analysis of the compatibility of current DSM-IV, proposed DSM-5.0, and DSM-5.1 diagnostic criteria in a correctional sample. Int J Offender Ther Comp Criminol, 2014. 58(6): p. 638–54.

31. McHugh, M.L., Interrater reliability: the kappa statistic. Biochem Med (Zagreb), 2012. 22(3): p. 276–82.

32. Landis, J.R. and G.G. Koch, The measurement of observer agreement for categorical data. Biometrics, 1977. 33(1): p. 159–74.

33. SAS Institute Inc. Base SAS ® 9.4 Procedures Guide Statistical Procedures Second Edition. Copyright © 2013. https://support.sas.com/documentation/cdl/en/procstat/66703/PDF/default/procstat.pdf.

34. Agrawal, A., A.C. Heath, and M.T. Lynskey, DSM-IV to DSM-5: the impact of proposed revisions on diagnosis of alcohol use disorders. Addiction, 2011. 106(11): p. 1935–43.

35. Mewton, L., T. Slade, O. McBride, R. Grove, and M. Teesson, An evaluation of the proposed DSM-5 alcohol use disorder criteria using Australian national data. Addiction, 2011. 106(5): p. 941–50.

36. Proctor, S.L., A.M. Kopak, and N.G. Hoffmann, Compatibility of current DSM-IV and proposed DSM-5 diagnostic criteria for cocaine use disorders. Addict Behav, 2012. 37(6): p. 722–8.

37. Kelly, S.M., J. Gryczynski, S.G. Mitchell, A. Kirk, K.E. O’Grady, and R.P. Schwartz, Concordance between DSM-5 and DSM-IV nicotine, alcohol, and cannabis use disorder diagnoses among pediatric patients. Drug Alcohol Depend, 2014. 140: p. 213–6.

38. Hasin, D., B.F. Grant, L. Cottler, J. Blaine, L. Towle, B. Ustun, and N. Sartorius, Nosological comparisons of alcohol and drug diagnoses: a multisite, multi-instrument international study. Drug Alcohol Depend, 1997. 47(3): p. 217–26.

39. Substance Abuse and Mental Health Services Administration. Resources for Screening, Brief Intervention, and Referral to Treatment (SBIRT). https://www.samhsa.gov/sbirt/resources

40. Saitz, R., Alcohol screening and brief intervention in primary care: Absence of evidence for efficacy in people with dependence or very heavy drinking. Drug Alcohol Rev, 2010. 29(6): p. 631–40.

